# Impact of Chronic Pain on the Families of U.S. Adults

**DOI:** 10.1101/2025.02.28.25322828

**Authors:** Jennifer S. De La Rosa, Katherine E. Herder, Rita D. Romero, De’Sha S. Wolf, Tally Largent-Milnes, Mohab M. Ibrahim, Stacy S. Pigott, Greg T. Chism, Beth E. Meyerson, Julie G. Pilitsis, Benjamin R. Brady, Allison J. Huff, Alicia M. Allen, Maria Manriquez, Kristyn Piñeda, Kyle A. Suhr, Taylor Young, Freya Spielberg, Todd W. Vanderah

## Abstract

Chronic pain (CP) can profoundly strain family systems, yet few population studies have explored CP with high impact on families. We used 2023 National Health Interview Survey Data (n=29,522) to characterize CP with high impact on families of U.S. adults. Findings are: (1) the prevalence of CP with high impact on families (HICP-Family) is 4.4% in U.S. adults and 18.1% in U.S. adults with CP; (2) HICP-Family is almost exclusively reported by those who have high impact chronic pain (HICP) that limits their individual functioning in life and work; (3) yet, among those with HICP, not all (46.3%) report HICP-Family; (4) HICP-Family can feasibly be modeled as a latter transition stage in a tiered cascade of advancing CP-associated impacts; (5) there is a strong association between clinically significant mental health symptoms and HICP-Family; HICP-Family is six times more prevalent among those who screen positive for moderate-to-severe anxiety or depression symptoms. Families highly impacted by CP may not be adequately resourced for adaptive functional resilience. HICP-Family may form an underappreciated barrier to achieving optimal pain and mental health outcomes for people with HICP. Further, without intervention, members of families highly impacted by CP may face heightened susceptibility to onset of suboptimal pain and mental health trajectories of their own. The need to develop effective supports and promote adaptive resilience in families highly impacted by CP is clear. Clinical tools to support person-centered assessment of family and relational functioning, effective family-level interventions, and innovative models of care are needed.

**IN BRIEF:** Establishes prevalence of chronic pain with high impact on families (HICP-Family); situates HICP-Family within emerging pain research frameworks.

## 1. INTRODUCTION

Chronic pain (CP) can profoundly impact family systems[75,76,96,97]. CP may challenge the family’s shared expectations[58,70,78,95,118] including who provides financial resources and emotional support [61,74], the conditions under which pain is considered a ‘legitimate’ rationale for functional limitations[14,75], and the acceptable forms, intensities, and frequencies of pain communications[33,48,49,116]. Research in mental health suggests the possibility of functional spillover effects as a mechanism for CP’s impact on families[71].

The gap between the family’s needs and their current reality greatly influences resilience to CP; mitigating factors can include the flexibility with which expectations are held, individual or interpersonal coping skills and supports, and the availability of financial and community resources promoting effective adaptation[30,41,42,52,63,66,97,119,123].

If the challenge presented by CP outmatches the family’s capacities and resources, relationships are likely to settle into dysfunctional patterns[4,38,42,97,113]. Families highly impacted by CP can be marked by cycles of emotional dysregulation and reactivity[16,17]; invalidation and stigma[33,129,130], catastrophic thinking[49]; frequent conflict[113]; role tension[2,115]; insufficient care and monitoring of children[126]; children acting as caregivers and surrogate parents [20,35,57,106,109,111]; dysfunctional regulation of relational closeness and distance[9,104,113,114]; overidentification with the subjective experiences of others[2,114]; infantilization of children and adults [84,98]; spousal and marital stress[62,74,122]; withdrawal from social networks and resultant isolation, loneliness, abuse or neglect[6,72,105]; and the suppression– or overly demonstrative expression of–anger, frustration, resentment, anxiety, despair, hopelessness, and pain[1,5,7,34,114].

Early pain socialization and family relational patterns are heavily intertwined: humans tend to express their pain to those with whom they feel relationally connected and from whom they anticipate the possibility of authentic experiences of care[2,18,30]. Maladaptive pain responses– overprotective/disempowering[19,21,40] or invalidating/dismissive[12,94]–to acute or chronic pain may frequently be associated with reduced or excessive family cohesion[93,107], marital dissatisfaction[74,92] and pain-attributed distress and disability[15]. Pain responses that effectively resolve the tension between acceptance and change may reduce pain intensity, [2,18,46] and build trust into the family system[60,133]. However, CP may also strain or upend previously positive patterns of communication and relational security[11,30]. Co-occurring mental health challenges may create a mutually reinforcing cycle with CP in families[8,11,25,26,31,73]. Family dysfunction is strongly predictive of pain-related disability and, like mental health, may confound the measurement of pain intensity[70,70,75,108].

Family members may blame themselves for being unable to soothe their loved ones’ pain[37]. They may feel that their supportive behaviors are not fully acknowledged nor reciprocated[3,65]; over time they may begin to experience burnout, resentfulness, and mistrust[137]. At the same time, people living with CP frequently report significant unmet needs for emotional validation and support and experience that they are being stigmatized, disbelieved, and isolated[94]. They may blame themselves for causing problems in the family and for the continuation of CP[22,69,88].

The clinical importance of CP’s impact on families is clear, yet few studies have explored population dynamics of CP with high impact on families (HICP-Family). We used 2023 National Health Interview Survey data to advance population-level understanding of HICP-Family in U.S. adults, aligning our analysis with emerging pain research frameworks that emphasize functional impact as a bridge between population and clinical studies[29,45,53,79,91,99,131].

## 2. METHODS

### 2.1 Study Design and Analysis Plan

We estimated the prevalence of U.S. adult chronic pain with high family impact as a proportion of the U.S. population and in millions of people. We evaluated the two-way interaction of chronic pain’s impact on individuals and chronic pain’s impact on families to assess whether family impact of pain is feasibly modeled as an advancement of functional impact beyond the individual. We then visualized the data as a cascade of functional impact in U.S. adults. Because high-impact pain in both individuals and families is so closely connected to mental health status[25,26,39], we estimated the prevalence of clinically significant anxiety and/or depression symptoms within each step in the CP-attributed functional impact cascade. Subsequently, we evaluated this relationship from the other direction, grouping the cascade by the presence/absence of clinically significant mental health symptoms a priori in order to examine and differences and commonalities between the two groups in terms of functional impact.

### 2.2 Data Source

We use data from the National Health Interview Survey (NHIS), a nationally representative, publicly available, deidentified data set suitable for calculating prevalence statistics about chronic pain in the noninstitutionalized U.S. adult population. The NHIS has been identified as the best data source for population-level surveillance of chronic pain[28]. Detailed information on the survey methodology has been published by the National Center for Health Statistics[90].

### 2.3 Measures

**Chronic Pain (CP)** was measured using the survey item *“In the past 3 months, how often did you have pain? Would you say never, some days, most days, or every day?”* Those who answered “Most days” or “Every day” were considered to have chronic pain, while those endorsing “Never” or “Some days” were considered not to have chronic pain. This is consistent with the International Association for the Study of Pain’s (IASP) definition of chronic pain as implemented in the International Classification of Disease (ICD-11)[121,136], and is the standard operationalization used by the National Center for Health Statistics.

**High-Impact Chronic Pain (HICP)** was measured using the survey item *“Over the past three months, how often did your pain limit your life or work activities? Would you say never, some days, most days, or every day?”* Those with chronic pain *and* who answered “Most days” or “Every Day” to this item were considered to have high-impact chronic pain (HICP); this is the standard operationalization used by the National Center for Health Statistics[77].

**Chronic Pain with high impact on Families (HICP-Family)** is a novel construct developed in this study. We operationalized HICP-Family using the survey item *“Over the past three months, how often did YOUR pain affect your family and significant others? Would you say never, some days, most days, or every day?”* For analysis of national prevalence (Tables 1 and 2), we considered those who answered “Most days” or “Every day” to this item to have chronic pain with high impact on families. For the analyses situating HICP-Family as part of CP-attributed functional impact cascade, the measurement of HICP-Family was constrained to U.S. adults who endorsed concomitant HCIP.

**Table 1.** Prevalence of chronic pain(CP) and CP-attributed functional impacts in millions of people and as a proportion of the United States adult population.

|  | U.S. Adult General Population |  |
| --- | --- | --- |
|  | N<br>[95 CI] | % of U.S. adults<br>[95 CI] |
| Chronic pain (CP) | 60.8 million<br>[58.5, 63.1] | 24.3%<br>[23.7, 25.0] |
| High-Impact CP (HICP) | 21.2 million<br>[20.0, 22.3] | 8.5%<br>[8.1, 8.9] |
| Chronic Pain with High Impact<br>on Family (HICP-Family) | 11.0 million [10.3, 11.8] | 4.4%<br>[4.1, 4.7] |

Data Source: National Center for Health Statistics, National Health Interview Survey, 2023 N = National prevalence in millions of US adults

**Table 2.** Two-way contingency table of high impact chronic pain (HICP) with chronic pain with high impact on families (HICP-Family), among the 60.8 million U.S. adults living with chronic pain.

|  |  | Chronic Pain with High Impact on Family |  |
| --- | --- | --- | --- |
|  |  | No | Yes |
| High Impact<br>Chronic Pain | No | 63.1% | 2.1% |
|  | Yes | 18.7% | 16.1% |

Data Source: National Center for Health Statistics, National Health Interview Survey, 2023

**Anxiety and/or Depression Symptoms (A/D)** were measured using two brief screeners for symptoms of anxiety and depression: 1) the General Anxiety Disorder-2 (GAD-2)[59], a validated brief scale used to screen for probable anxiety, and (2) the Patient Health Questionnaire-2 (PHQ-2)[67], a validated brief scale used to screen for probable depression. We operationalized positive screening results in line with the standard cut-points. Respondents who scored 3 or higher on the GAD-2 were considered to have anxiety symptoms[59]; respondents who scored 3 or higher on the PHQ-2 were considered to have depressive symptoms[68]. Survey respondents were coded as having anxiety and/or depression symptoms (A/D) if they scored greater than or equal to 3 on the PHQ-2 the GAD-2, or both.

### 2.4 Missing Data

Responses recorded as “Refused,” “Don’t Know,” or “Not Ascertained” in responses to questions used for the above measures were considered missing data. Missing data for questions used to operationalize chronic pain CP, HICP, HICP-Family, and A/D included a total of 1101 observations, representing 3.7% of overall data. Complete case analysis was used.

### 2.5 Analysis Details

Population prevalence and population mean estimates presented in all tables and figures were calculated using SAS statistical software version 9.4 accounting for the stratification, clustering, and weighting procedures of the complex NHIS survey design.

## 3. RESULTS

An estimated 4.4% [95% CI: 4.1, 4.7] of the U.S. adult population*—*18.1% of U.S. adults with CP*—*report HICP-Family **(Table 1).**

If high impact of chronic pain on families could be feasibly conceptualized as an advancement in pain’s impact spilling over from the individual and into the family system, we would expect to see few respondents endorsing HICP-Family who did not concomitantly endorse HICP. We explored the interaction of HICP and HICP-Family **(Table 2).** Just 2.1% of U.S. adults with chronic pain endorsed high family impact without also endorsing HICP; the small amount of non-conforming data suggest the empirical feasibility of modeling HICP-Family as an advancement of chronic pain’s functional impact beyond the directly affected individual.

Next, we tabulated and visualized model-conformant data (100% - 2.1% = 97.1% of data was conformant) as a tiered “cascade” of advancing functional impact **(Table 3**, **Figure 1)**. The presence of high-impact pain did not deterministically predict high family impact: fewer than half (46.3% [95% CI: 44.1%, 48.5%]) of U.S. adults with high-impact chronic pain endorsed high family impact. Among all those who endorsed high family impact of chronic pain, 88.5% [95% CI: 86.4%, 90.6%] concomitantly endorsed HICP.

**Table 3:**
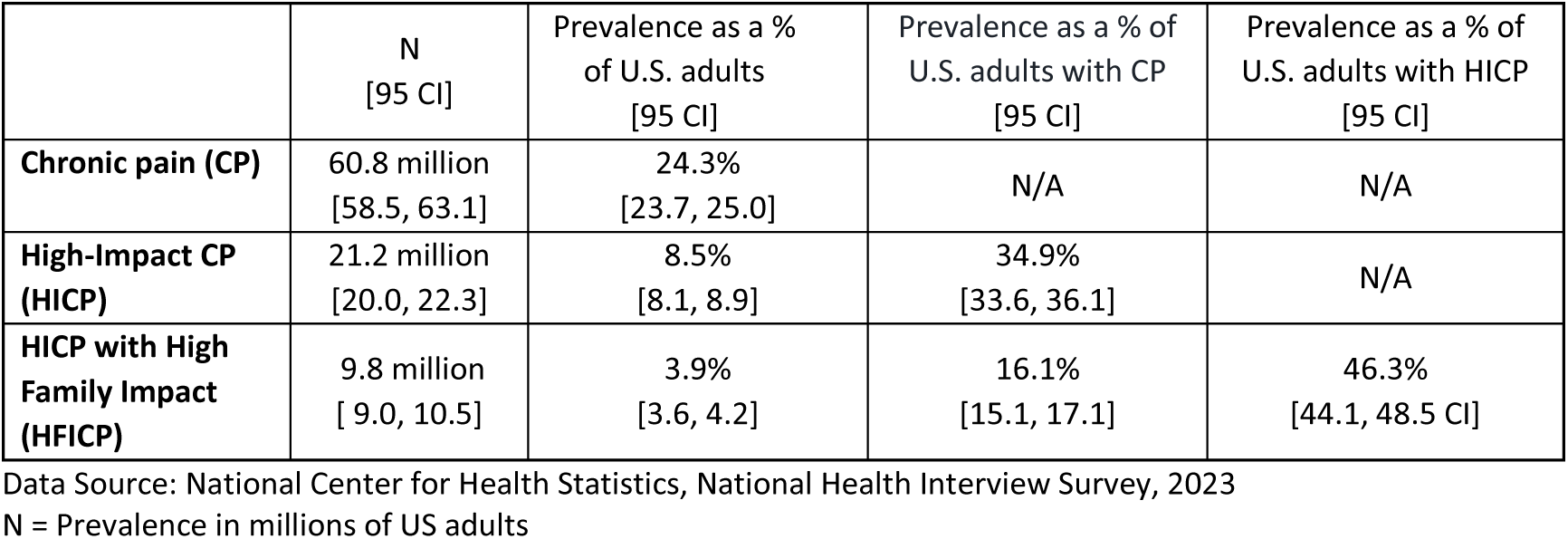
Impact of chronic pain on U.S. adults and their families: a tiered cascade of advancing functional impact.

|  | N<br>[95 CI] | Prevalence as a %<br>of U.S. adults<br>[95 CI] | Prevalence as a % of<br>U.S. adults with CP<br>[95 CI] | Prevalence as a % of<br>U.S. adults with HICP<br>[95 CI] |
| --- | --- | --- | --- | --- |
| Chronic pain (CP) | 60.8 million<br>[58.5, 63.1] | 24.3%<br>[23.7, 25.0] | N/A | N/A |
| High-Impact CP<br>(HICP) | 21.2 million<br>[20.0, 22.3] | 8.5%<br>[8.1, 8.9] | 34.9%<br>[33.6, 36.1] | N/A |
| HICP with High<br>Family Impact<br>(HFICP) | 9.8 million<br>[ 9.0, 10.5] | 3.9%<br>[3.6, 4.2] | 16.1%<br>[15.1, 17.1] | 46.3%<br>[44.1, 48.5 CI] |

N = Prevalence in millions of US adults

**Figure 1.**
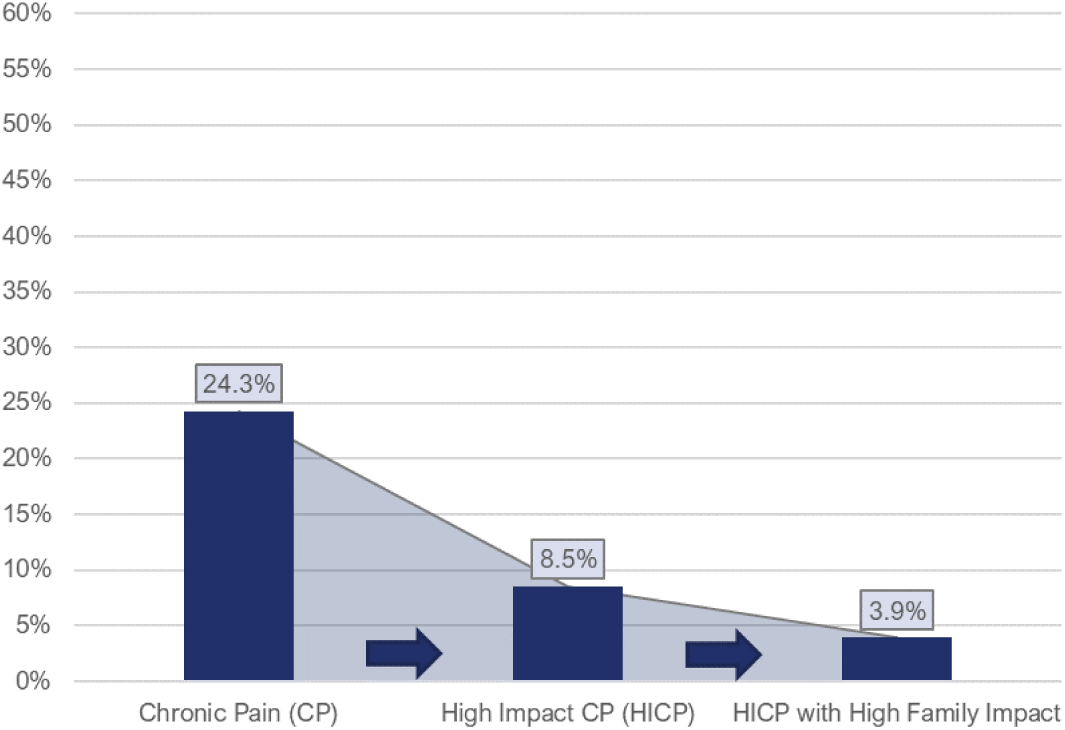
Prevalence of chronic pain (CP) and associated impacts in U.S. adults: a tiered cascade of advancing functional impact Data Source: National Center for Health Statistics, National Health Interview Survey, 2023

Because the literature so closely links the impact of chronic pain with mental health status[25,26,39], we estimated the prevalence of clinically significant anxiety/depression symptoms within each step in the functional impact cascade **(Table 4):**

**Table 4.** Prevalence of clinically significant anxiety/depression symptoms modeled as a tiered cascade of CP-attributed functional impact.

|  | Prevalence of A/D symptoms (%) |
| --- | --- |
| **U.S. adults** | 12.0% [11.5%, 12.5%] |
| **U.S. adults with Chronic Pain (CP)** | 24.4% [23.1%, 25.6%] |
| **U.S. adults with high-impact CP (HICP)** | 37.1% [34.8%, 39.5%] |
| **U.S. adults with HICP and high impact of CP on family (HICP-Family)** | 46.1% [42.6%, 49.6%] |

To further assess the intersection of A/D with CP, HICP, and HICP-Family, we separated the functional impact cascade into two groups based on positive/negative screening for clinically significant anxiety and depression symptoms (**Figure 2**, **Table 5**). The prevalence of chronic pain and CP-attributed impact on individuals and families is markedly elevated in those who screened positive for A/D symptoms compared to others. The magnitude of this disparity increases exponentially as functional impact advances: CP is (49.0%/20.9%) =2.35 times more prevalent in those with A/D. HICP is (26.1%/6.0%) = 4.35 times more prevalent in those with A/D; and family impact of pain is (14.9%/2.4%) = 6.20 times more prevalent in those with A/D (**Figure 2**, **Table 5**).

**Figure 2.**
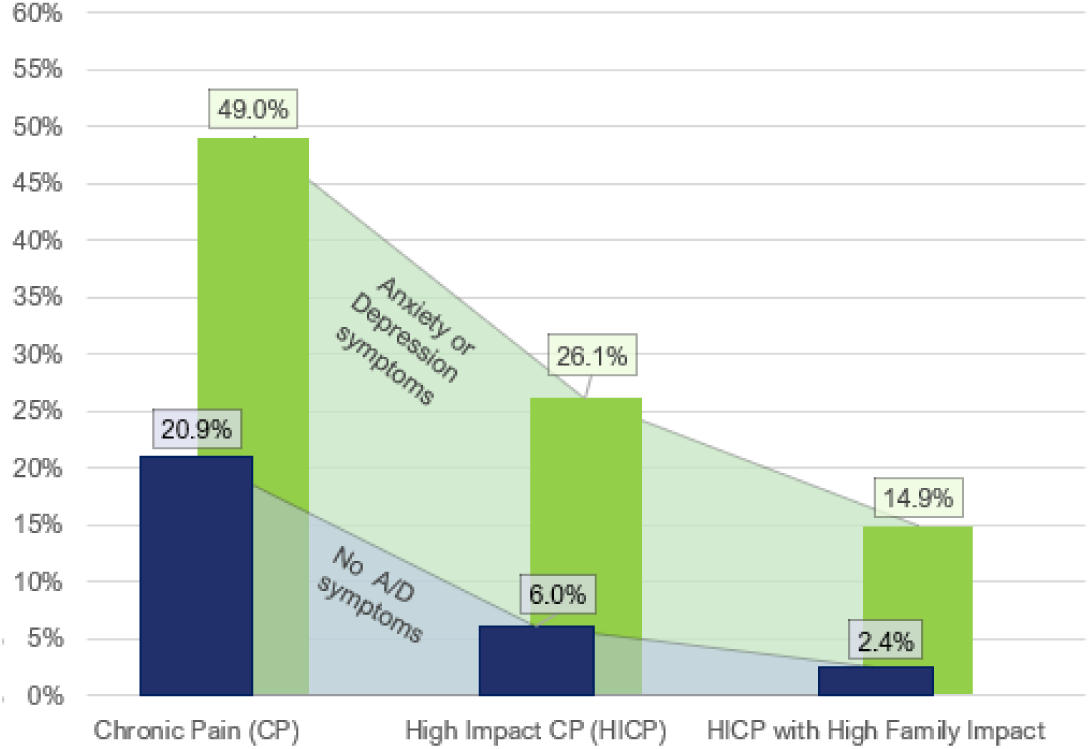
Prevalence of chronic pain (CP) and associated impacts as a proportion of U.S. adults: modeled as a tiered cascade of advancing functional impacts, grouped by presence of clinically significant anxiety/depression symptoms Data Source: National Center for Health Statistics, National Health Interview Survey, 2023

**Table 5.** Prevalence of chronic pain (CP) and associated impacts as a proportion of U.S. adults: modeled as a tiered cascade of advancing functional impacts, grouped by presence of clinically significant anxiety/depression symptoms.

|  | U.S. Adults without Anxiety/Depression Symptoms |  |  |  | U.S. Adults with Anxiety/ Depression Symptoms |  |  |  |
| --- | --- | --- | --- | --- | --- | --- | --- | --- |
|  | N<br>[95 CI] | Prevalence<br>as a % of<br>U.S. adults<br>[95 CI] | Prevalence<br>as a % of<br>U.S. adults<br>with CP<br>[95 CI] | Prevalence<br>as a % of<br>U.S. adults<br>with HICP<br>[95 CI] | N<br>[95 CI] | Prevalence<br>as a % of<br>U.S. adults<br>[95 CI] | Prevalence<br>as a % of<br>U.S. adults<br>with CP<br>[95 CI] | Prevalence<br>as a % of U.S.<br>adults with<br>HICP<br>[95 CI] |
| **Chronic pain** | 45.6 million<br>[43.8, 47.5] | 20.9%<br>[20.2, 21.5] | N/A | N/A | 14.7 million<br>[13.7, 15.7] | 49.0%<br>[46.9, 51.2] | N/A | N/A |
| **High-impact chronic pain (HICP)** | 13.2 million<br>[12.4, 14.0] | 6.0%<br>[5.7, 6.4] | 28.9%<br>[27.6, 30.3] | N/A | 7.8 million<br>[7.1, 8.5] | 26.1%<br>[24.2, 27.9] | 53.1%<br>[50.3, 56.0] | N/A |
| **HICP with High Impact on Family (HICP-Family)** | 5.2 million<br>[4.7, 5.7] | 2.4%<br>[2.2, 2.6] | 11.4%<br>[10.5, 12.4] | 39.6%<br>[36.9, 42.3] | 4.5 million<br>[4.0, 4.9] | 14.9%<br>[13.5, 16.3] | 30.5% [27.9, 33.0] | 57.5%<br>[53.7, 61.2] |

Data Source: National Center for Health Statistics, National Health Interview Survey, 2023
N = National prevalence in millions of US adults

## 4. DISCUSSION

Pain is mainly communicated and attended to in families; families are a primary building block of communities; community-level disparities in chronic pain may likely be reinforced and maintained through its unequal concentration in families, across geographies and generations. Health conditions associated with chronic pain may run in families biologically and/or environmentally, such that families experience repeated burden of similar types of pain at similar points in the life course[117]. Family systems already highly impacted by CP may be under-resourced to provide caregiving and support to family members; evidence suggests that these family members may become especially susceptible to onset of their own suboptimal pain and mental health trajectories[24,55,55,86,117,124].

The degree to which the chronic pain of U.S. adults impacts their family systems has not been well characterized. The present study represents an initial step in advancing population-level understanding. We estimated that 11 million U.S. adults, 4.4% of the U.S. adult population, report chronic pain that affects their families most days or every day (HICP-Family) U.S. adults whose chronic pain has high impact on their family make up 18.1% of all U.S. adults living with chronic pain**. (Table 1).**

We found no empirical difficulty in aligning HICP-Family with the field’s general effort to capture and catalogue dynamic temporal transitions in pain’s functional impact. HICP could be conceptualized as a necessary, but not sufficient, condition*—*fewer than half (46.3%) of U.S. adults with HICP endorsed high family impact **(Figure 1**, **Table 3)**. The data are consistent with conceptualizing HICP-Family as a distinct transition in CP impact: representing unmitigated advancement of CP-attributed functional limitations in the individual and creating spillover impacts on family systems **(Table 2)**. Finally, we documented (bidirectionally) the association between anxiety/depression and chronic pain’s functional impact cascade. Clinically significant anxiety and depression symptoms were increasingly prevalent at each subsequent step in the chronic pain impact cascade: among all U.S. adults, the prevalence of clinically significant anxiety/depression symptoms is 12.0%. In U.S. adults with CP, 24.4% have A/D symptoms; among U.S. adults with HICP, 37.1% have A/D symptoms; and among U.S. adults with HICP-Family, 46.1% have A/D symptoms **(Table 4)**. Upon reversing the contingency and comparing the functional cascade among those with A/D to those without A/D, we found that the prevalence of chronic pain and downstream functional impact is markedly higher in those with anxiety/depression symptoms compared to others and that the magnitude of the disparity between those with and without A/D increases exponentially as functional impact increases **(Figure 2**, **Table 5).**

Despite calls for family-oriented approaches to the treatment of chronic pain by leaders in the field[96], relatively little research has addressed family functioning. This gap is particularly striking in the context of adult chronic pain. Validated, person-centered, culturally resonant approaches to the assessment of family (and individual) functioning are needed[43]. Measurement of family functioning will not be an easy task: chronic pain can have complicated, significant, and widespread impacts across family systems, and it does not restrict its impact to direct caregivers of people living with pain. Chronic pain impacting those who would otherwise be involved in providing care reduces the family’s *collective* caregiving resources. In this way, family dependents who are not affected by pain, such as children, older family members, and those with disabilities, may suffer lack of care, due to a family member’s reduced capacity to provide it. High-impact parental pain can advance profound adversity on children; these children’s experiences are similar to those reported by children of parents with mental illness and/or substance use disorders[124,125]. The siblings of children with high-impact pain may experience that parents/guardians are distracted and emotionally taxed; the entire family may become primarily focused on the needs of the child with pain. Grandparents and other extended family members who have high-impact pain may be limited in their social engagement and caregiving to children. Family members who do not require caregiving can suffer as well: Pain may put enormous financial and emotional stress on spousal partners and others who pick up the slack created by reductions in the productive capacity of the family unit. Mid-life caregivers of elders with high-impact chronic pain may often be pulled between providing care for the generation above and the generation below, particularly in communities where elder caregiving is culturally expected[44]. In some cases, family conflict related to chronic pain may catalyze estrangement. Providing care for people with high-impact pain may not be as health-promoting as receiving it. It is possible that caregivers of people with chronic pain neglect their own health to focus on their family member. Finally, many families will have more than one individual living with high-impact chronic pain. Research to address family impact of pain will need to grapple with these complexities.

In pain research, family members are often categorized with a binary valence: as a resource promoting resilience, or as a negative influence that may require being “worked around”, clinically. In addition to focusing on attributes (positive and negative) of the individuals and observing pain-related interactions, emerging research exploring the jointly held interactional quality and networked structure of relationships –including but not limited to the context of pain[13,100,127,128,133,134] –may be a productive research direction moving forward.

Outcomes for both chronic pain and mental health are heavily associated with perceived availability of social support[132]. Involving families in treatment, particularly in communities with heavy burden of chronic pain and CP-attributed impacts, may be a more efficacious approach compared to treatment as usual–which considers patients primarily as individuals[13,51,80,81]. Interventions provided by behavioral health peer and family recovery support specialists may offer an opportunity to develop family-systems interventions and natural supports[82,83], particularly in communities affected by shortages of behavioral health care providers. Increased engagement of psychosocial and occupational rehabilitation professionals in multidisciplinary pain care could also be beneficial. As a non-limiting example, occupational therapists may often place *less* emphasis on a joint’s range of motion, and *more* emphasis on the adaptive resilience of a human’s daily life, leveraging patients’ innate drive for mastery [50,110,112]. Adaptive recovery of meaningful, self-directed lives is critical to the recovery and sustained resilience of individuals and their families; the effective and adaptive functioning of family systems could become a high-value endpoint for future research.

## 5. LIMITATIONS

Several limitations are noted. First, persons with no permanent address, persons living on military bases, U.S. expatriates, and individuals residing in long-term care institutions, and correctional facilities are not represented in the NHIS. Second, the 2023 NHIS adult dataset included only the brief forms of the screeners for depression and anxiety; though these have been shown to be viable alternatives to the GAD7 and PHQ-8[56], it is still possible that using the brief form could in some way influence the measurement of anxiety/depression. The NHIS measures the impact of the respondent’s pain on the family, as perceived by the respondent, however, not all individuals may easily recognize the impact of their pain on their family, which could lead to undercounting the true prevalence. Ideally, information on family impact would be collected from family members themselves. Last, contextual information such as how many family members are affected by a respondent’s pain, whether they see themselves as caregivers, care receivers, or some other relationship, whether they currently cohabitate with the respondent, and whether respondents have experienced CP-associated family estrangement, are unavailable in the NHIS public use files, limiting disentangling of cohabitation versus kinship and preventing a more nuanced analysis of the subjective meaning respondents attributed to the survey item concerning family impact of pain.

## 6. CONCLUSION

> *“Try imagining a place where it’s always safe and warm / Come in, she said / I’ll give ya, shelter from the storm”* --Bob Dylan

Chronic pain is indeed a ‘sensitive barometer of population health’[135] that is disproportionately prevalent in under-resourced and stigmatized individuals, families, and communities[10,64,102,103,120,135]. It is perhaps less widely understood that among individuals living with chronic pain, vulnerability to high-impact functional limitations *also* exists along a social, cultural, relational, and psychological gradient[23,27,32,36,47,54,77,85,87,89,101]. We suggest that HICP-Family could play an underrecognized role in maintaining feedback loops of anxiety/depression, functional limitations, and chronic pain.

More research is needed to understand transitions in functional impact: including the predictors of transitions from CP to HICP, and the transition from HICP to HICP-Family. To better understand HICP-Family, tools supporting person-centered clinical assessment of family and relational functioning are needed. Investment is needed to learn what can be done to prevent or reverse the spillover effects of CP-associated functional limitations (HICP) on family systems. Helping CP-compromised families to recover represents a meaningful opportunity in improving quality of life for millions of children, adults, and older people who are directly and indirectly affected by high-impact chronic pain, and potentially in addressing recalcitrant disparities across clinical and sociodemographic subpopulations.

## Data Availability

No original datasets were generated over the course of this study, all data used in this study are publicly available at:
https://www.cdc.gov/nchs/nhis/documentation/2023-nhis.html

## 7. ACKNOWLEDGEMENTS

Many on our co-author team have had important experiences with their own or a family member’s pain. Our professional activity as clinicians, scientists, and communicators is usefully informed by this experiential expertise. We acknowledge Sally W. Schultz, PhD, OTR, LPC, and Professor Emerita at Texas Woman’s University, and Doreen DeRoss, LPC, for several helpful conversations informing our thinking on pain and families. Funding for this study was provided by the Comprehensive Center for Pain & Addiction at the University of Arizona Health Sciences. The authors have no conflicts of interest to declare. No original data sets were generated over the course of this study; the data used in this study are publicly available on the National Health Interview Survey website.

## REFERENCES

[1] Aaron RV, Fisher EA, de la Vega R, Lumley MA, Palermo TM. Alexithymia in individuals with chronic pain and its relation to pain intensity, physical interference, depression, and anxiety: a systematic review and meta-analysis. PAIN 2019;160:994. doi:10.1097/j.pain.0000000000001487.

[2] Akbari F, Mohammadi S, Dehghani M, Sanderman R, Hagedoorn M. Interpretations of partners’ responses to pain behaviours: Perspectives of patients and partners. Br J Health Psychol 2021;26:401–418. doi:10.1111/bjhp.12490.

[3] Akbari R, Farsi Z, Sajadi SA. Relationship between fatigue and quality of life and related factors in family caregivers of patients on hemodialysis. BMC Psychiatry 2023;23:430. doi:10.1186/s12888-023-04934-2.

[4] Ashton-James CE, Ziadni MS. Uncovering and Resolving Social Conflicts Contributing to Chronic Pain: Emotional Awareness and Expression Therapy. J Health Serv Psychol 2020;46:133–140. doi:10.1007/s42843-020-00017-y.

[5] Ballús-Creus C, Rangel MV, Peñarroya A, Pérez J, Leff J. Expressed emotion among relatives of chronic pain patients, the interaction between relatives’ behaviours and patients’ pain experience. Int J Soc Psychiatry 2014;60:197–205. doi:10.1177/0020764013496371.

[6] Bannon S, Greenberg J, Mace RA, Locascio JJ, Vranceanu A-M. The role of social isolation in physical and emotional outcomes among patients with chronic pain. Gen Hosp Psychiatry 2021;69:50–54.

[7] Beutler LE, Daldrup R, Engle D, Guest P, Corbishley A, Meredith KE. Family dynamics and emotional expression among patients with chronic pain and depression. PAIN 1988;32:65. doi:10.1016/0304-3959(88)90024-3.

[8] Beveridge JK, Noel M, Soltani S, Neville A, Orr SL, Madigan S, Birnie KA. The association between parent mental health and pediatric chronic pain: a systematic review and meta-analysis. PAIN 2024;165:997. doi:10.1097/j.pain.0000000000003125.

[9] Block AR, Kremer EF, Gaylor M. Behavioral treatment of chronic pain: The spouse as a discriminative cue for pain behavior. PAIN 1980;9:243. doi:10.1016/0304-3959(80)90011-1.

[10] Booker SQ. African Americans’ Perceptions of Pain and Pain Management: A Systematic Review. J Transcult Nurs 2016;27:73–80. doi:10.1177/1043659614526250.

[11] Boone D, Kim SY. Family Strain, Depression, and Somatic Amplification in Adults with Chronic Pain. Int J Behav Med 2019;26:427–436.

[12] Boring BL, Walsh KT, Nanavaty N, Mathur VA. Shame Mediates the Relationship Between Pain Invalidation and Depression. Front Psychol 2021;12:743584. doi:10.3389/fpsyg.2021.743584.

[13] Breiger RL. The duality of persons and groups. Soc Forces 1974;53:181–190. Available: https://academic.oup.com/sf/article-abstract/53/2/181/2229911. Accessed 22 Feb 2025.

[14] Burns JW, Gerhart J, Post KM, Smith DA, Porter LS, Buvanendran A, Fras AM, Keefe FJ. Spouse Criticism/Hostility Toward Partners With Chronic Pain: The Role of Spouse Attributions for Patient Control Over Pain Behaviors. J Pain 2018;19:1308–1317. doi:10.1016/j.jpain.2018.05.007.

[15] Campbell L, DiLorenzo M, Atkinson N, Riddell RP. Systematic Review: A Systematic Review of the Interrelationships Among Children’s Coping Responses, Children’s Coping Outcomes, and Parent Cognitive-Affective, Behavioral, and Contextual Variables in the Needle-Related Procedures Context. J Pediatr Psychol 2017;42:611–621.

[16] Campbell P, Jordan KP, Smith BH, Scotland G, Dunn KM. Chronic pain in families: a cross-sectional study of shared social, behavioural, and environmental influences. PAIN 2018;159:41. doi:10.1097/j.pain.0000000000001062.

[17] Cano A, Miller LR, Loree A. Spouse Beliefs about Partner Chronic Pain. J Pain Off J Am Pain Soc 2009;10:486–492. doi:10.1016/j.jpain.2008.11.005.

[18] Cano A, Williams AC de C. Social interaction in pain: Reinforcing pain behaviors or building intimacy? Pain 2010;149:9–11. doi:10.1016/j.pain.2009.10.010.

[19] Chambers CT, Craig KD, Bennett SM. The impact of maternal behavior on children’s pain experiences: an experimental analysis. J Pediatr Psychol 2002;27:293–301.

[20] Chikhradze N, Knecht C, Metzing S. Young carers: growing up with chronic illness in the family - a systematic review 2007-2017. J Compassionate Health Care 2017;4:12. doi:10.1186/s40639-017-0041-3.

[21] Clementi MA, Faraji P, Cordts KP, MacDougall K, Wilson A, Palermo TM, Holley AL. Parent Factors are Associated with Pain and Activity Limitations in Youth with Acute Musculoskeletal Pain: A Cohort Study. Clin J Pain 2019;35:222–228. doi:10.1097/AJP.0000000000000668.

[22] Coady A, Godard R, Holtzman S. Understanding the link between pain invalidation and depressive symptoms: The role of shame and social support in people with chronic pain. J Health Psychol 2024;29:52–64. doi:10.1177/13591053231191919.

[23] Cook BL, Zuvekas SH, Carson N, Wayne GF, Vesper A, McGuire TG. Assessing Racial/Ethnic Disparities in Treatment across Episodes of Mental Health Care. Health Serv Res 2014;49:206–229. doi:10.1111/1475-6773.12095.

[24] Dario AB, Kamper SJ, O’Keeffe M, Zadro J, Lee H, Wolfenden L, Williams CM. Family history of pain and risk of musculoskeletal pain in children and adolescents: a systematic review and meta-analysis. PAIN 2019. doi:10.1097/j.pain.0000000000001639.

[25] De La Rosa JS, Brady BR, Herder KE, Wallace JS, Ibrahim MM, Allen AM, Meyerson BE, Suhr KA, Vanderah TW. The unmet mental health needs of U.S. adults living with chronic pain. PAIN 2024:10.1097/j.pain.0000000000003340. doi:10.1097/j.pain.0000000000003340.

[26] De La Rosa JS, Brady BR, Ibrahim MM, Herder KE, Wallace JS, Padilla AR, Vanderah TW. Co-occurrence of chronic pain and anxiety/depression symptoms in U.S. adults: prevalence, functional impacts, and opportunities. PAIN 2023:10.1097/j.pain.0000000000003056. doi:10.1097/j.pain.0000000000003056.

[27] Derricks V, Hirsh AT, Perkins AJ, Daggy JK, Matthias MS. Health Care Discrimination Affects Patient Activation, Communication Self-Efficacy, and Pain for Black Americans. J Pain 2024;25:104663. Available: https://www.sciencedirect.com/science/article/pii/S1526590024006199. Accessed 22 Feb 2025.

[28] Duca LM, Helmick CG, Barbour KE, Nahin RL, Von Korff M, Murphy LB, Theis K, Guglielmo D, Dahlhamer J, Porter L, Falasinnu T, Mackey S. A Review of Potential National Chronic Pain Surveillance Systems in the United States. J Pain 2022;23:1492–1509.

[29] Eccleston C, Begley E, Birkinshaw H, Choy E, Crombez G, Fisher E, Gibby A, Gooberman-Hill R, Grieve S, Guest A, Jordan A, Lilywhite A, Macfarlane GJ, McCabe C, McBeth J, Pickering AE, Pincus T, Sallis HM, Stone S, Van der Windt D, Vitali D, Wainwright E, Wilkinson C, de C Williams AC, Zeyen A, Keogh E. The establishment, maintenance, and adaptation of high- and low-impact chronic pain: a framework for biopsychosocial pain research. Pain 2023;164:2143–2147.

[30] Eccleston C, Crombez G, Scotford A, Clinch J, Connell H. Adolescent chronic pain: patterns and predictors of emotional distress in adolescents with chronic pain and their parents. Pain 2004;108:221–229.

[31] Eccleston C, Wastell S, Crombez G, Jordan A. Adolescent social development and chronic pain. Eur J Pain 2008;12:765–774. doi:10.1016/j.ejpain.2007.11.002.

[32] Edmond SN, Heapy AA, Kerns RD. Engaging Mental Health Professionals in Addressing Pain. JAMA Psychiatry 2019;76:565–566.

[33] Edmond SN, Keefe FJ. Validating pain communication: current state of the science. Pain 2015;156:215–219. Available: https://journals.lww.com/pain/fulltext/2015/02000/validating_pain_communication_current_state_of.4.aspx. Accessed 26 Jan 2025.

[34] Ellis JD, Resko SM, Brown S, Agius E, Kollin R, Burlaka V. Correlates of Expressed Emotion Among Family Members of Individuals Who Sought Treatment for Opioid Use. J Nerv Ment Dis 2020;208:870. doi:10.1097/NMD.0000000000001220.

[35] Evans R. Children as Caregivers. In: Ben-Arieh A, Casas F, Frønes I, Korbin JE, editors. Handbook of Child Well-Being: Theories, Methods and Policies in Global Perspective. Dordrecht: Springer Netherlands, 2014. pp. 1893–1916. doi:10.1007/978-90-481-9063-8_173.

[36] Falasinnu T, Hossain MB, Weber KA, Helmick CG, Karim ME, Mackey S. The Problem of Pain in the United States: A Population-Based Characterization of Biopsychosocial Correlates of High Impact Chronic Pain Using the National Health Interview Survey. J Pain 2023;24:1094–1103.

[37] Fales JL, Essner BS, Harris MA, Palermo TM. When Helping Hurts: Miscarried Helping in Families of Youth With Chronic Pain. J Pediatr Psychol 2014;39:427–437. doi:10.1093/jpepsy/jsu003.

[38] Fang Y, Liu M, Wu M, Liu M, Niu T, Zhang X. Pain intensity and self-perceived burden mediate the relationship between family functioning and pain catastrophizing in patients with neuropathic pain. Nurs Health Sci 2024;26:e13097. doi:10.1111/nhs.13097.

[39] Fang Y, Liu M, Wu M, Zhang J, Liu M, Niu T, Zhang X. Path Analysis Between Family Functioning and Mental Health in People With Neuropathic Pain: Roles of Pain Intensity, Self-Perceived Burden, Pain Catastrophizing, and Functional Status. Pain Manag Nurs 2024;25:e287–e294. doi:10.1016/j.pmn.2024.03.014.

[40] Fang Y, Shi L, Qin F, Li T, Zhang X, Li M. Influence of Family-Learned Fear-of-Pain on Patients. Pain Manag Nurs 2024;25:11–18. doi:10.1016/j.pmn.2023.04.003.

[41] Flink IK, Reme S, Jacobsen HB, Glombiewski J, Vlaeyen JWS, Nicholas MK, Main CJ, Peters M, Williams AC de C, Schrooten MGS, Shaw W, Boersma K. Pain psychology in the 21st century: lessons learned and moving forward. Scand J Pain 2020;20:229–238. doi:10.1515/sjpain-2019-0180.

[42] Flor H, Turk DC, Scholz OB. Impact of chronic pain on the spouse: marital, emotional and physical consequences. J Psychosom Res 1987;31:63–71.

[43] Freynhagen R, Fullen BM, Reneman MF, Treede R-D. Functioning in chronic pain: a call for a global definition. PAIN 2024;165:2235. doi:10.1097/j.pain.0000000000003307.

[44] Gallant MP, Spitze G, Grove JG. Chronic Illness Self-care and the Family Lives of Older Adults: A Synthetic Review Across Four Ethnic Groups. J Cross-Cult Gerontol 2010;25:21–43. doi:10.1007/s10823-010-9112-z.

[45] Gatchel RJ, Reuben DB, Dagenais S, Turk DC, Chou R, Hershey AD, Hicks GE, Licciardone JC, Horn SD. Research Agenda for the Prevention of Pain and Its Impact: Report of the Work Group on the Prevention of Acute and Chronic Pain of the Federal Pain Research Strategy. J Pain 2018;19:837–851. doi:10.1016/j.jpain.2018.02.015.

[46] Gauthier N, Thibault P, Sullivan MJL. Individual and relational correlates of pain-related empathic accuracy in spouses of chronic pain patients. Clin J Pain 2008;24:669–677.

[47] George SZ, Allen KD, Alvarez C, Arbeeva L, Callahan LF, Nelson AE, Schwartz TA, Golightly YM. Prevalence and Factors Associated With High-Impact Chronic Pain in Knee Osteoarthritis: The Johnston County Health Study. J Pain 2024;25:104687. doi:10.1016/j.jpain.2024.104687.

[48] Goubert L, Bernardes SF. Interpersonal dynamics in chronic pain: The role of partner behaviors and interactions in chronic pain adjustment. Curr Opin Psychol 2025:101997. doi:10.1016/j.copsyc.2025.101997.

[49] Goubert L, Eccleston C, Vervoort T, Jordan A, Crombez G. Parental catastrophizing about their child’s pain. The parent version of the Pain Catastrophizing Scale (PCS-P): A preliminary validation. PAIN 2006;123:254. doi:10.1016/j.pain.2006.02.035.

[50] Grajo LC. Occupational Adaptation as a Normative and Intervention Process: New Perspectives on Schkade and Schultz’s Professional Legacy. Adaptation Through Occupation: Multidimensional Perspectives. Taylor and Francis, 2024. pp. 83–104. doi:10.4324/9781003522508-8.

[51] Green HD, Pescosolido BA. Social pathways to care: how community-based network ties shape the health care response of individuals with mental health problems. Soc Psychiatry Psychiatr Epidemiol 2024;59:431–442. doi:10.1007/s00127-023-02476-2.

[52] Hassett AL, Finan PH. The Role of Resilience in the Clinical Management of Chronic Pain. Curr Pain Headache Rep 2016;20:39. doi:10.1007/s11916-016-0567-7.

[53] Hayes C, Hodson FJ. A Whole-Person Model of Care for Persistent Pain: From Conceptual Framework to Practical Application. Pain Med 2011;12:1738–1749. doi:10.1111/j.1526-4637.2011.01267.x.

[54] Herman PM, Broten N, Lavelle TA, Sorbero ME, Coulter ID. Exploring the prevalence and construct validity of high-impact chronic pain across chronic low-back pain study samples. Spine J Off J North Am Spine Soc 2019;19:1369–1377.

[55] Higgins KS, Birnie KA, Chambers CT, Wilson AC, Caes L, Clark AJ, Lynch M, Stinson J, Campbell-Yeo M. Offspring of parents with chronic pain: a systematic review and meta-analysis of pain, health, psychological, and family outcomes. PAIN 2015;156:2256. doi:10.1097/j.pain.0000000000000293.

[56] Hlynsson JI, Carlbring P. Diagnostic accuracy and clinical utility of the PHQ-2 and GAD-2: a comparison with long-format measures for depression and anxiety. Front Psychol 2024;15. doi:10.3389/fpsyg.2024.1259997.

[57] How caregiving affects development: Psychological implications for child, adolescent, and adult caregivers. Washington, DC, US: American Psychological Association, 2009.

[58] Hudson M, Johnson MI. Hidden family rules: perspective on a dysfunctional paternalistic system and the persistence of pain. Front Pain Res 2023;4:1303853. doi:10.3389/fpain.2023.1303853.

[59] Hughes AJ, Dunn KM, Chaffee T, Bhattarai J (Jackie), Beier M. Diagnostic and clinical utility of the GAD-2 for screening anxiety symptoms in individuals with multiple sclerosis. Arch Phys Med Rehabil 2018;99:2045–2049. doi:10.1016/j.apmr.2018.05.029.

[60] Issner JB, Cano A, Leonard MT, Williams AM. How do I empathize with you? Let me count the ways: Relations between facets of pain-related empathy. J Pain 2012;13:167–175. doi:10.1016/j.jpain.2011.10.009.

[61] Kemler MA, Furnée CA. The Impact of Chronic Pain on Life in the Household. J Pain Symptom Manage 2002;23:433–441. doi:10.1016/S0885-3924(02)00386-X.

[62] Kerns RD, Turk DC. Depression and Chronic Pain: The Mediating Role of the Spouse. J Marriage Fam 1984;46:845–852. doi:10.2307/352532.

[63] Kerns RD, Weiss LH. Family Influences on the Course of Chronic Illness: a Cognitive-Behavioral Transactional Model. Ann Behav Med 1994;16:116–121. doi:10.1093/abm/16.2.116.

[64] Khalatbari-Soltani S, Blyth FM. Socioeconomic position and pain: a topical review. PAIN 2022;163:1855. doi:10.1097/j.pain.0000000000002634.

[65] Kindt S, Vansteenkiste M, Loeys T, Cano A, Lauwerier E, Verhofstadt LL, Goubert L. When Is Helping your Partner with Chronic Pain a Burden? The Relation Between Helping Motivation and Personal and Relational Functioning. Pain Med 2015;16:1732–1744. doi:10.1111/pme.12766.

[66] Kinnie KR, Vance DE, Patrician PA, Billings R, Aroke EN. Chronic Pain Resilience Across Clinical Populations: A Concept Analysis. Pain Manag Nurs 2024;25:442–450. doi:10.1016/j.pmn.2024.03.019.

[67] Kroenke K, Spitzer RL, Williams JBW. The Patient Health Questionnaire-2: Validity of a Two-Item Depression Screener. Med Care 2003;41:1284. doi:10.1097/01.MLR.0000093487.78664.3C.

[68] Kroenke K, Spitzer RL, Williams JBW. The Patient Health Questionnaire-2: validity of a two-item depression screener. Med Care 2003;41:1284–1292.

[69] Kuharic M, Sharp LK, Turpin RS, Mulhern B, Lee TA, Grace Rose CE, Monteiro A, Pickard AS. Care recipient self-perceived burden: Perspectives of individuals with chronic health conditions or personal experiences with caregiving on caregiver burden in the US. SSM - Qual Res Health 2024;5:100398. doi:10.1016/j.ssmqr.2024.100398.

[70] Langford DJ, Lou R, Sheen S, Amtmann D, Colloca L, Edwards RR, Farrar JT, Katz NP, McDermott MP, Reeve BB. Expectations for improvement: a neglected but potentially important covariate or moderator for chronic pain clinical trials. J Pain 2023;24:575–581. Available: https://www.sciencedirect.com/science/article/pii/S1526590022004795. Accessed 26 Jan 2025.

[71] Lee D, Kim Y, Devine B. Spillover Effects of Mental Health Disorders on Family Members’ Health-Related Quality of Life: Evidence from a US Sample. Med Decis Making 2022;42:80–93. doi:10.1177/0272989X211027146.

[72] Lee S, Small BJ, Cawthon PM, Stone KL, Almeida DM. Social activity diversity as a lifestyle factor to alleviate loneliness and chronic pain. J Psychosom Res 2023;172:111434. doi:10.1016/j.jpsychores.2023.111434.

[73] Leonard MT, Cano A. Pain affects spouses too: Personal experience with pain and catastrophizing as correlates of spouse distress. PAIN 2006;126:139. doi:10.1016/j.pain.2006.06.022.

[74] Leonard MT, Cano A, Johansen AB. Chronic Pain in a Couples Context: A Review and Integration of Theoretical Models and Empirical Evidence. J Pain 2006;7:377–390. doi:10.1016/j.jpain.2006.01.442.

[75] Lewandowski AS, Palermo TM, Stinson J, Handley S, Chambers CT. Systematic review of family functioning in families of children and adolescents with chronic pain. J Pain Off J Am Pain Soc 2010;11:1027–1038. doi:10.1016/j.jpain.2010.04.005.

[76] Lewandowski W, Morris R, Draucker CB, Risko J. Chronic Pain and the Family: Theory-Driven Treatment Approaches. Issues Ment Health Nurs 2007;28:1019–1044. doi:10.1080/01612840701522200.

[77] Lucas JW, Sohi I. Chronic Pain and High-impact Chronic Pain in U.S. Adults, 2023. 2024.

[78] Lynch-Jordan AM, Kashikar-Zuck S, Szabova A, Goldschneider KR. The Interplay of Parent and Adolescent Catastrophizing and Its Impact on Adolescents’ Pain, Functioning, and Pain Behavior. Clin J Pain 2013;29:681–688. doi:10.1097/AJP.0b013e3182757720.

[79] Macgregor C, Walumbe J, Tulle E, Seenan C, Blane DN. Intersectionality as a theoretical framework for researching health inequities in chronic pain. Br J Pain 2023;17:479–490. doi:10.1177/20494637231188583.

[80] Martire LM. The “Relative” Efficacy of Involving Family in Psychosocial Interventions for Chronic Illness: Are There Added Benefits to Patients and Family Members? Fam Syst Health 2005;23:312–328.

[81] Martire LM, Schulz R. Involving Family in Psychosocial Interventions for Chronic Illness. Curr Dir Psychol Sci 2007;16:90–94. doi:10.1111/j.1467-8721.2007.00482.x.

[82] Matthias MS, Adams JM, Eliacin J. Volunteer Peer Support for Chronic Pain Self-Management: A Qualitative Study of Benefits and Barriers. J Gen Intern Med 2024. doi:10.1007/s11606-024-09217-4.

[83] Matthias MS, Bair MJ, Ofner S, Heisler M, Kukla M, McGuire AB, Adams J, Kempf C, Pierce E, Menen T, McCalley S, Johnson NL, Daggy J. Peer Support for Self-Management of Chronic Pain: the Evaluation of a Peer Coach-Led Intervention to Improve Pain Symptoms (ECLIPSE) Trial. J Gen Intern Med 2020;35:3525–3533. doi:10.1007/s11606-020-06007-6.

[84] Meldrum ML, Tsao JC-I, Zeltzer LK. “I Can’t Be What I Want to Be”: Children’s Narratives of Chronic Pain Experiences and Treatment Outcomes. Pain Med 2009;10:1018–1034. doi:10.1111/j.1526-4637.2009.00650.x.

[85] Miró J, Roman-Juan J, Sánchez-Rodríguez E, Solé E, Castarlenas E, Jensen MP. Chronic Pain and High Impact Chronic Pain in Children and Adolescents: A Cross-Sectional Study. J Pain 2023;24:812–823. doi:10.1016/j.jpain.2022.12.007.

[86] Monin JK, Schulz R. Interpersonal effects of suffering in older adult caregiving relationships. Psychol Aging 2009;24:681–695.

[87] Mun CJ, Davis MC, Molton IR, Karoly P, Suk HW, Ehde DM, Tennen H, Kerns RD, Jensen MP. Personal resource profiles of individuals with chronic pain: Sociodemographic and pain interference differences. Rehabil Psychol 2019;64:245. Available: https://psycnet.apa.org/record/2019-04367-001. Accessed 22 Feb 2025.

[88] Muñoz-Peña IJ, González-Gutiérrez JL, Yunta-Rua L, Pacho-Hernández JC, López-López A. Stress, perceived competence and guilt as predictors of depression in parents with chronic pain. Front Psychol 2025;15. doi:10.3389/fpsyg.2024.1473955.

[89] Nahin RL, Feinberg T, Kapos FP, Terman GW. Estimated Rates of Incident and Persistent Chronic Pain Among US Adults, 2019-2020. JAMA Netw Open 2023;6:e2313563. doi:10.1001/jamanetworkopen.2023.13563.

[90] National Center for Health Statistics. National Health Interview Survey Description. 2024 Available: https://ftp.cdc.gov/pub/Health_Statistics/NCHS/Dataset_Documentation/NHIS/2023/srvydesc-508.pdf. Accessed 7 Jul 2023.

[91] Nelson SM, Cunningham NR, Kashikar-Zuck S. A Conceptual Framework for Understanding the Role of… : The Clinical Journal of Pain. n.d. Available: https://journals.lww.com/clinicalpain/FullText/2017/03000/A_Conceptual_Framework_for_Understanding_the_Role.10.aspx. Accessed 3 Feb 2025.

[92] Newton-John TR, Williams AC de C. Chronic pain couples: perceived marital interactions and pain behaviours. Pain 2006;123:53–63.

[93] Nicassio PM, Radojevic V. Models of family functioning and their contribution to patient outcomes in chronic pain. Motiv Emot 1993;17:295–316. doi:10.1007/BF00992224.

[94] Nicola M, Correia H, Ditchburn G, Drummond P. Invalidation of chronic pain: a thematic analysis of pain narratives. Disabil Rehabil 2021;43:861–869. doi:10.1080/09638288.2019.1636888.

[95] Palermo TM. Impact of recurrent and chronic pain on child and family daily functioning: a critical review of the literature. J Dev Behav Pediatr JDBP 2000;21:58–69.

[96] Palermo TM, Chambers CT. Parent and family factors in pediatric chronic pain and disability: an integrative approach. Pain 2005;119:1–4.

[97] Palermo TM, Valrie CR, Karlson CW. Family and parent influences on pediatric chronic pain: A developmental perspective. Am Psychol 2014;69:142–152.

[98] Pao M, Bosk A. Anxiety in Medically Ill Children/Adolescents. Depress Anxiety 2011;28:40–49. doi:10.1002/da.20727.

[99] Patel M, Johnson AJ, Booker SQ, Bartley EJ, Palit S, Powell-Roach K, Terry EL, Fullwood D, DeMonte L, Mickle AM, Sibille KT. Applying the NIA Health Disparities Research Framework to Identify Needs and Opportunities in Chronic Musculoskeletal Pain Research. J Pain 2022;23:25–44. doi:10.1016/j.jpain.2021.06.015.

[100] Perry BL, Pescosolido BA, Small ML, McCranie A. Introduction to the Special Issue on Ego Networks. Netw Sci 2020;8:137–141. doi:10.1017/nws.2020.18.

[101] Pitcher MH, Von Korff M, Bushnell MC, Porter L. Prevalence and Profile of High-Impact Chronic Pain in the United States. J Pain 2019;20:146–160. doi:10.1016/j.jpain.2018.07.006.

[102] Raghuraman N, Akintola T, Rassu FR, O’Connor TD, Chen S, Gruber-Baldini A, Colloca L. The Effects of Socioeconomic Position on Endogenous Pain Modulation: A Quasi-Experimental Approach. J Pain 2025:104778. doi:10.1016/j.jpain.2025.104778.

[103] Rassu FS, McFadden M, Aaron RV, Wegener ST, Ephraim PL, Lane E, Brennan G, Minick KI, Fritz JM, Skolasky RL. The Relationship Between Neighborhood Deprivation and Perceived Changes for Pain-Related Experiences Among US Patients with Chronic Low Back Pain During the COVID-19 Pandemic. Pain Med 2021;22:2550–2565. doi:10.1093/pm/pnab179.

[104] Riffin C, Fried T, Pillemer K. Impact of Pain on Family Members and Caregivers of Geriatric Patients. Clin Geriatr Med 2016;32:663–675. doi:10.1016/j.cger.2016.06.010.

[105] Rokach A, Rosenstreich E, Brill S, Goor Aryeh I. People with Chronic Pain and Caregivers: Experiencing Loneliness and Coping with It. Curr Psychol 2018;37:886–893. doi:10.1007/s12144-017-9571-2.

[106] Rolland JS. Parental illness and disability: a family systems framework. J Fam Ther 1999;21:242–266. doi:10.1111/1467-6427.00118.

[107] Romano JM, Turner JA, Jensen MP. The Family Environment in Chronic Pain Patients: Comparison to Controls and Relationship to Patient Functioning. J Clin Psychol Med Settings 1997;4:383–395. doi:10.1023/A:1026253418543.

[108] Saito T, Shibata M, Hirabayashi N, Honda T, Morisaki Y, Anno K, Sudo N, Hosoi M, Ninomiya T. Family dysfunction is associated with chronic pain in a community-dwelling Japanese population: The Hisayama study. Eur J Pain Lond Engl 2023;27:518–529.

[109] Saragosa M, Frew M, Hahn-Goldberg S, Orchanian-Cheff A, Abrams H, Okrainec K. The Young Carers’ Journey: A Systematic Review and Meta Ethnography. Int J Environ Res Public Health 2022;19:5826. doi:10.3390/ijerph19105826.

[110] Schkade JK, Schultz S. Occupational adaptation: Toward a holistic approach for contemporary practice: I. Am J Occup Ther 1992;46:829–837.

[111] Schorr S, Goldner L. “Like stepping on glass”: A theoretical model to understand the emotional experience of childhood parentification. Fam Relat 2023;72:3029–3048. doi:10.1111/fare.12833.

[112] Schultz S, Schkade JK. Occupational Adaptation: Toward a Holistic Approach for Contemporary Practice, Part 2. Am J Occup Ther 1992;46:917–925. doi:10.5014/ajot.46.10.917.

[113] Schwartz L, Slater MA, Birchler GR. The role of pain behaviors in the modulation of marital conflict in chronic pain couples. PAIN 1996;65:227. doi:10.1016/0304-3959(95)00211-1.

[114] Smith AA, Friedemann M-L. Perceived family dynamics of persons with chronic pain. J Adv Nurs 1999;30:543–551. doi:10.1046/j.1365-2648.1999.01123.x.

[115] Snelling J. The effect of chronic pain on the family unit. J Adv Nurs 1994;19:543–551.

[116] Stephens MAP, Martire LM, Cremeans-Smith JK, Druley JA, Wojno WC. Older women with osteoarthritis and their caregiving husbands: Effects of pain and pain expression on husbands’ well-being and support. Rehabil Psychol 2006;51:3–12.

[117] Stone AL, Wilson AC. Transmission of risk from parents with chronic pain to offspring: an integrative conceptual model. PAIN 2016;157:2628. doi:10.1097/j.pain.0000000000000637.

[118] Strunin L, Boden LI. Family consequences of chronic back pain. Soc Sci Med 1982 2004;58:1385–1393.

[119] Sturgeon JA, Zubieta C, Kaplan CM, Pierce J, Arewasikporn A, Slepian PM, Hassett AL, Trost Z. Broadening the Scope of Resilience in Chronic Pain: Methods, Social Context, and Development. Curr Rheumatol Rep 2024;26:112–123. doi:10.1007/s11926-024-01133-0.

[120] Topping M, Fletcher J. Educational attainment, family background and the emergence of pain gradients in adulthood. Soc Sci Med 1982 2024;346:116692.

[121] Treede R-D, Rief W, Barke A, Aziz Q, Bennett MI, Benoliel R, Cohen M, Evers S, Finnerup NB, First MB, Giamberardino MA, Kaasa S, Korwisi B, Kosek E, Lavand’homme P, Nicholas M, Perrot S, Scholz J, Schug S, Smith BH, Svensson P, Vlaeyen JWS, Wang S-J. Chronic pain as a symptom or a disease: the IASP Classification of Chronic Pain for the International Classification of Diseases (ICD-11). Pain 2019;160:19–27.

[122] Turk DC, Flor H, Rudy TE. Pain and families. I. Etiology, maintenance, and psychosocial impact. Pain 1987;30:3–27.

[123] Turk DC, Gatchel RJ. Psychological Approaches to Pain Management, Second Edition: A Practitioner’s Handbook. Guilford Publications, 2013.

[124] Umberger W. Children of Parents With Chronic Noncancer Pain: A Comprehensive Review of the Literature. J Child Adolesc Psychiatr Nurs Off Publ Assoc Child Adolesc Psychiatr Nurses Inc 2014;27:26–34.

[125] Umberger W, Martsolf D, Jacobson A, Risko J, Patterson M, Calabro M. The shroud: ways adolescents manage living with parental chronic pain. J Nurs Scholarsh Off Publ Sigma Theta Tau Int Honor Soc Nurs 2013;45:344–354.

[126] Umberger WA, Risko J, Covington E. The forgotten ones: challenges and needs of children living with disabling parental chronic pain. J Pediatr Nurs 2015;30:498–507.

[127] Van Alboom M, Baert F, Bernardes SF, Verhofstadt L, Bracke P, Jia M, Musial K, Gabrys B, Goubert L. Examining the Role of Structural and Functional Social Network Characteristics in the Context of Chronic Pain: An Ego-centered Network Design. J Pain 2024;25:104525.

[128] Van Alboom M, Elmer T, Boersma K, Forgeron P, Baert F, Bracke P, Goubert L. Social integration of adolescents with chronic pain: a social network analysis. PAIN 2022;163:2232. doi:10.1097/j.pain.0000000000002623.

[129] Wakefield EO, Belamkar V, Litt MD, Puhl RM, Zempsky WT. “There’s Nothing Wrong With You”: Pain-Related Stigma in Adolescents With Chronic Pain. J Pediatr Psychol 2021;47:456–468.

[130] Wallwork SB, Shenk C, McMurtry CM, Hood AM, Pavlova M, Olson AE, Moseley GL, Noel M. “I hear you”. Validation in the context of children’s pain as an untapped opportunity to prevent chronic pain. Pain 2024;165:2667–2672.

[131] Wideman TH, Edwards RR, Walton DM, Martel MO, Hudon A, Seminowicz DA. The Multimodal Assessment Model of Pain: A Novel Framework for Further Integrating the Subjective Pain Experience Within Research and Practice. Clin J Pain 2019;35:212. doi:10.1097/AJP.0000000000000670.

[132] Wilson JM, Colebaugh CA, Meints SM, Flowers KM, Edwards RR, Schreiber KL. Loneliness and Pain Catastrophizing Among Individuals with Chronic Pain: The Mediating Role of Depression. J Pain Res 2022;15:2939–2948. doi:10.2147/JPR.S377789.

[133] Woods SB, Roberson PNE, Booker Q, Wood BL, Booker SQ. Longitudinal Associations of Family Relationship Quality With Chronic Pain Incidence and Persistence Among Aging African Americans. J Gerontol Ser B 2024;79:gbae064. doi:10.1093/geronb/gbae064.

[134] Yang B, Anderson Z, Zhou Z, Liu S, Haase CM, Qu Y. The Unique and Interactive Roles of Neural Reward Sensitivity and Family Conflict in Predicting Youth’s Internalizing Problems: A Biopsychosocial Approach. Psychoneuroendocrinology 2023;153:106253. doi:10.1016/j.psyneuen.2023.106253.

[135] Zajacova A, Grol-Prokopczyk H, Zimmer Z. Sociology of Chronic Pain. J Health Soc Behav 2021;62:302–317. doi:10.1177/00221465211025962.

[136] Zelaya CE, Dahlhamer JM, Lucas JW, Connor, Eric M. Chronic pain and high-impact chronic pain among U.S. adults, 2019. NCHS Data Brief No 390 2020. Available: https://stacks.cdc.gov/view/cdc/97308. Accessed 28 Mar 2024.

[137] Zhao H, Kulbok PA, Williams IC, Manning C, Logan JG, Romo RD. Exploring Experiences of Pain Management Among Family Caregivers of Community-Dwelling Older Adults With Dementia. Am J Hosp Palliat Med 2024;41:927–933. doi:10.1177/10499091231210290.

